# Drug company methodologies used for reporting in the UK pharmaceutical industry payment transparency database between 2015-2019: A content analysis

**DOI:** 10.1101/2024.04.12.24305382

**Authors:** James Larkin, Britta Matthes, Mohamed Azribi, Conor Kearns, Shai Mulinari, Emily Rickard, Frank Moriarty, Tom Fahey, Piotr Ozieranski

## Abstract

**Background:** Pharmaceutical companies make hundreds of millions of pounds in marketing/R&D-related payments annually to healthcare organisations and healthcare professionals. UK pharmaceutical industry self-regulatory bodies require member companies who sign up to their code of conduct to publish details of their payments. They are also required to publish the methodologies underlying these payments - methodological notes. This study aimed to analyse UK pharmaceutical companies’ methodological notes and their adherence to the relevant codes of conduct and guidance.

**Methods:** We conducted a content analysis of methodological notes for the years 2015, 2017 and 2019 and assessed companies’ adherence to self-regulatory bodies’ requirements and recommendations for methodology disclosure.

**Results:** Overall, 90 companies made payment disclosures in all three years, publishing 269 methodological notes. We found gaps in adherence to self-regulatory requirements. Only 3 (3.3%) companies provided clear information for all self-regulatory body recommendations and regulations in all of their notes. Companies also varied in their approaches to important areas. For example, of the 244 notes with clear information on VAT management, 36.1% (N=88) included VAT, 30.3% (N=74) excluded VAT, and 33.6% (N=82) had multiple rules for VAT.

**Conclusions:** There was evidence of widespread non-adherence to self-regulatory requirements. This suggests flaws with self-regulation and a need for greater enforcement of rules or consideration of a publicly mandated disclosure system.

## 1. Introduction

The UK pharmaceutical industry pays healthcare professionals (HCPs) and healthcare organisations (HCOs) hundreds of millions of pounds annually for research, consultancy services, education initiatives, collaborations, and other activities.^1,2^ These payments can affect health policy and practice.^3–5^ While industry actors argue that these payments are instrumental in drug development and marketing,^6,7^ there is evidence that they can create a conflict of interest (COI) where a HCO or HCP’s primary interest in patient care conflicts with the secondary interest of financial gain.^3^ Numerous studies have outlined how such payments can negatively affect prescribing,^4,5^ publishing,^8^ clinical guideline recommendations^9^ and other areas.^10,11^ Furthermore, some of these payments have been found to serve as inducements to prescribe or procure medicines, notably in the United States (US),^12^ Greece,^13^ Italy,^14^ and Croatia.^14^ In response to such practices (and other issues such as fraud), some countries (e.g. the US, France and Portugal) have legally mandated that companies disclose details of some of these payments.^15–17^ In other jurisdictions (e.g. the UK, Ireland), disclosure is governed by self-regulatory systems managed by pharmaceutical industry trade groups.^17,18^

Although payment disclosure has been presented as a key step in addressing COIs,^19,20^ international evidence suggests that some opacity remains.^15,17,21,22^ In countries which follow self-regulatory approaches to disclosure, issues exist with the structure of payment disclosure databases, their user-friendliness, and data presentation.^1,2,17,18,22^ This can limit data trustworthiness and one’s ability to identify COIs, which is important as the payment information is of interest to patients,^23,24^ journalists,^25^ policy-makers,^26^ and academics.^1^

The UK disclosure system is considered the most accessible industry-run system in Europe.^22,27^ The Association of the British Pharmaceutical Industry (ABPI) requires that member companies publish details of their payment methodologies (‘methodological notes’), alongside details of payments, every year on its website Disclosure UK (full overview of Disclosure UK in Box 1).^28^ ^22,27^ The ABPI represent the vast majority of UK manufacturers of branded prescription pharmaceuticals^28^ and in 2019 on Disclosure UK 130 companies disclosed £542.5 million in payments to HCPs and HCOs.^29^ Issues have been highlighted with the UK disclosure system, for example, the misreporting/underreporting of payments.^1^

Methodological notes are expected to provide necessary technical explanations for the published payment data in each country (e.g. whether VAT is included in a disclosure).^30^ Consideration of these explanations is necessary for building valid interpretations of the payment data.^31^ Any variations in disclosure practice are significant as they can undermine the validity and reliability of the data.

To date, methodological notes in the UK and elsewhere remain understudied. A notable exception is an Irish study which found that companies’ practices varied in many ways, for example, whether they include VAT or payments related to over-the-counter-medicines in their disclosures.^18^ Such inconsistencies create several issues,^32–34^ for example, making it difficult to compare companies’ payment values.^32^ However, the Irish study^18^ and others^1,18,35^ of methodological notes have been limited in scope, only analysing a limited number of years and a limited number of aspects of the notes.^1,18,35^

This study aimed to comprehensively analyse the content of methodological notes of companies reporting payments on the ABPI payment disclosure website for 2015-2019 and their adherence to the relevant codes of conduct and guidance. Specifically, we aimed to assess a) whether methodologies varied between companies and years, b) whether methodologies adhered to the ABPI code of practice and other regulatory advice, and c) what methodologies companies used in key areas (e.g. VAT and exchange rates).

Overall, evaluating methodological notes provides insight into the extent to which individual companies are complying with the rules set out by the trade groups,^28^ and therefore provides insights into self-regulation more broadly.

## 2. Methods

This is an observational study of three years of methodological notes: 2015, 2017 and 2019, for companies reporting payments on the ABPI’s Disclosure UK website.

### 2.1 Data collection

Methodological notes were downloaded in June 2017 (2015 data), December 2020 (2017 data) and July 2022 (2019 data) from the Disclosure UK website. Given that a primary aim of the study was to assess whether methodologies varied between years, we excluded notes of companies which did not report payments in all three years.

#### Box 1. Overview of UK Disclosure System

Companies which have signed up to the ABPI code of conduct are required to report relevant payments to healthcare professionals (HCPs), healthcare organisations (HCOs) and other relevant decision makers (ORDMs) on a centralised website: Disclosure UK.^28^ Payments are reported annually.^28^ Along with this, companies are required to upload ‘methodological notes’ alongside each annual disclosure of payments. Payment data and the associated methodological notes are removed from Disclosure UK three years after initial upload (e.g. 2019 data is published in June 2020 is removed in June 2023).^28^ Disclosed payments cover the following areas: contracted services (previously referred to as fee for service and consultancy), contributions to cost related to events, donations and grants to HCOs, research and development, and joint working, (renamed collaborative working in 2021, this payment category is unique to the UK, involving projects with contributions from both drug companies and HCOs).^28^ Full ABPI definitions for HCPs, HCOs and ORDMs and all of the above payment categories can be found in Appendix, eBox 1.

The ABPI disclosure system is part of the European Federation of Pharmaceutical Industries and Associations (EFPIA) Code of Practice (or “EFPIA Code”) and disclosure processes, similar to those described above, exist in several other European countries.

#### 2.1.1 Data extraction and coding framework

A proforma was developed (by PO) in Microsoft Excel to facilitate data extraction. The proforma was refined and revised iteratively based on feedback (from BM, ER, MA, CK and JL). The proforma consisted of headings based on requirements from the ABPI code of conduct,^36,37^ guidance from the Prescription Medicines Code of Practice Authority (PMCPA)^38,39^ – the ABPI’s self-regulatory authority administering the ABPI code,^40^ and, areas of interest from previous research.^18^

The ABPI requires companies to include details in their methodological note of how VAT, exchange rates, multi-year contracts and timing are handled.^36,37^ Timing refers to the methodology used to assign a date to a payment disclosure. For example, if an event occurs in 2014 but payment occurs in 2015, companies need to explain whether the payment is disclosed in the event year or payment year.

The PMCPA lists areas that ‘may be helpful when preparing methodological notes’.^38,39^ These include: cross-border payments, payments related to over-the-counter-medicines and payments related to medical devices.^38,39^ ^38,39^ For cross-border payments there were two distinct areas: 1) payments to UK registered recipients by non-UK affiliates and 2) payments to recipients registered outside the UK by the UK pharmaceutical company. The PMCPA also lists ‘other aspects for consideration’ in methodological notes.^38,39^ These include: ‘Non-monetary payments’ and ‘Going beyond minimum requirements’.^39^ We interpreted ‘Going beyond minimum requirements’ as any payment area that was included in disclosures without being required by the ABPI.

Additional variables, not included in the PMCPA guidance or ABPI code, were included based on previous research examining methodological notes:^18^

1. partial disclosure: whether HCPs are permitted to anonymise some payments they have received from a company and reveal their identity for other payments
2. non-attendances/cancellation: whether unrecouped payments for events or activities that do not occur or are not attended are disclosed
3. blinded market research: whether blinded market research payments are not disclosed or are reported in aggregate
4. internal events: whether any payments are disclosed for events organised by the company for HCPs
5. notable exclusions: payments areas not reported despite no explicit permission in ABPI guidance for that exclusion
6. notable aggregations: payments areas reported in aggregate despite no permission in ABPI guidance for aggregation

Each methodological note was searched by one of five researchers (BM, ER, MA, CK or JL) for data related to each area. Search terms were developed for each heading with further terms added during data extraction. If relevant information was found, it was pasted under the relevant heading in the proforma.

### 2.2 Data analysis

For each heading in the proforma there were two subheadings, a) information provided and b) methodology. For *information provided*, there were four categorisations: 1) Clear information provided, 2) Information provided but unclear, 3) Information not provided and 4) Described by the company as not applicable to their disclosure. These categories were deductively coded by JL. Examples of the categorisation for *clear information provided* and *information provided but unclear* are in Appendix, eTable 1.

The *methodology* was coded using conventional (i.e., inductive) content analysis to develop categorisations of methodologies.^41^ One of five researchers (JL, BM, ER, CK, PO) read through the relevant information from all companies for an individual area (e.g. VAT) and developed codes to describe companies’ methodologies in that area in a series of author meetings. For example, for VAT, coders developed three methodology categories: 1) VAT included in disclosures, 2) VAT excluded and 3) different rules depending on circumstances. Once coding was completed, a subheading entitled ‘change of methodology’ was added to capture if, for a given area (e.g. VAT), the methodology was different in at least two years. This subheading only applied to companies which had provided information on an area in at least two years. One researcher (JL) checked all coding and a second researcher (BM) reviewed 10% of all entries. Any discrepancies were discussed and resolved by JL and BM. Percentages and frequencies were used to analyse the data for both companies and notes. The primary analysis focuses on notes, and company level analysis is available in Appendix, eBox 2.

## 3. Results

Overall, 90 companies disclosed payments in 2015, 2017 and 2019 and were hence included for analysis. This represents 59.2% of the 152 companies which disclosed payments in at least one of the three years, and 95% of payments in terms of value for the three years. One company, Alimera Sciences, had not published a methodological note in 2017 (despite disclosing payments for this year), therefore, 269 methodological notes were available for analysis. Eleven companies (12.2%) published one note each in a format with reduced accessibility (e.g. image), and 72.7% (N=8) of these were 2015 notes. In 2017, two companies (2.2%) republished their 2015 methodological notes.

### 3.1 Provision of Information

Figure 1 details the percentage of notes adhering to ABPI requirements and PMCPA recommendations and provision of information in other areas (full details in Appendix, eTable 2).

**Figure 1.**
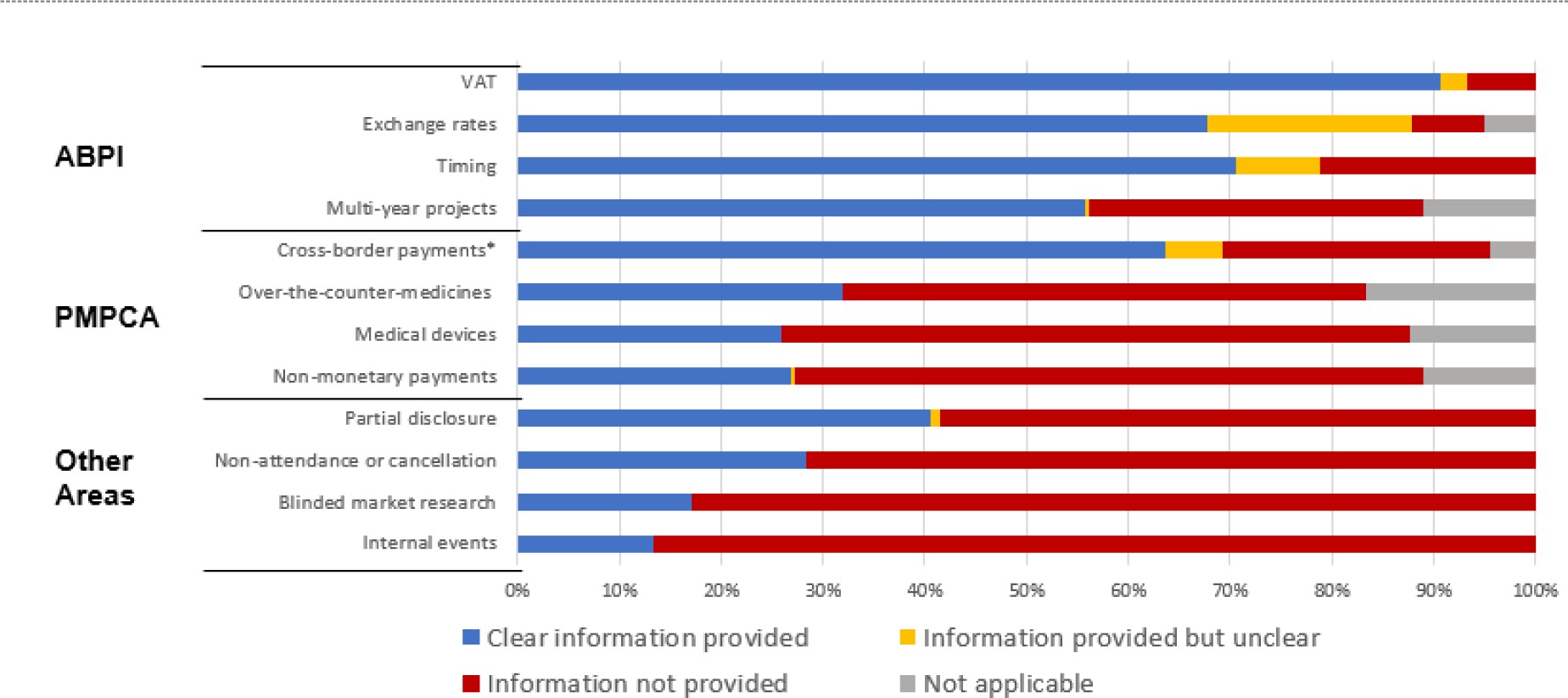
Provision of information for regulatory requirements/recommendations. Note: Not applicable means that this area was described by the company as not applicable to their disclosure in the given year. *For cross-border payments information was coded as “clear” if clear information was provided in relation to payments to UK registered recipients or in relation to non-UK registered recipients.

Compliance with ABPI requirements varied. For example, for 6.7% (N=18) of methodological notes, clear information on VAT management was not provided. For 32.7% (N=88) of notes, clear information was not provided on management of multi-year projects (note: in all cases where it is stated that clear information was not provided, this also means that clear information was not provided on whether this area was not applicable). Compliance with PMCPA recommendations also varied depending on the area but tended to be lower than compliance with ABPI requirements (see Figure 1 for comparisons). For 26.4% (N=71) of notes, clear information was not provided on management of cross-border payments. While 61.7% (N=166) of notes, did not include information on whether payments related to medical devices were covered by disclosures. When comparing across companies, 8.9% (N=8) of companies did not provide clear information in at least one of their methodological notes for all eight ABPI requirements/PMCPA recommendations. The median number of ABPI requirements/PMCPA recommendations that companies did not provide clear information in at least one of their methodological notes for was 3 (IQR:2-5). Only three (3.3%) companies provided clear information for all PMCPA recommendations and ABPI regulations in all of their notes (full details in Figure 2).

**Figure 2.**
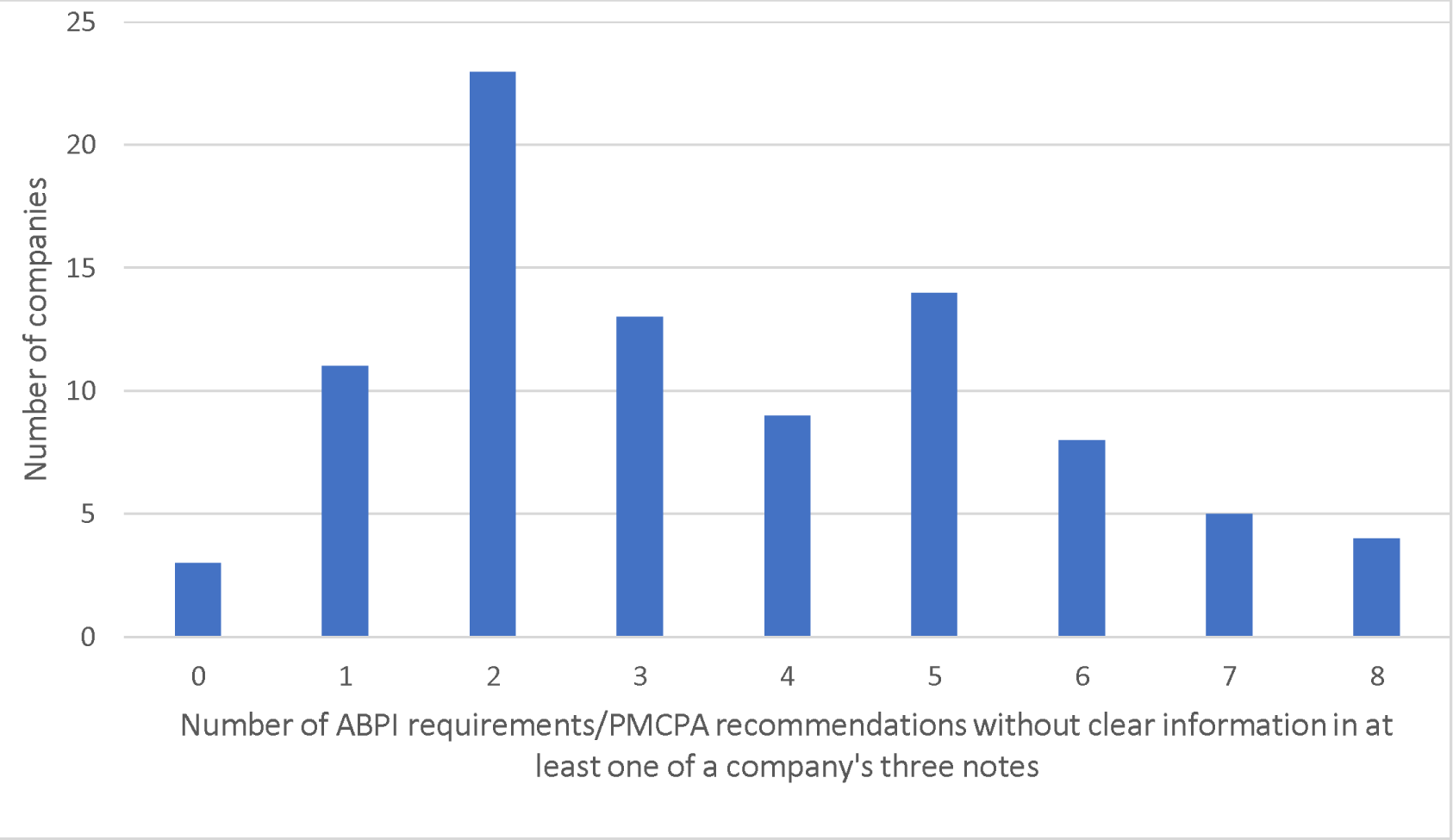
Companies not providing clear information for regulatory requirements/recommendations.

### 3.2 Methodologies for ABPI Requirements

With regard to ABPI requirements, companies’ methodologies varied. Table 1 provides an overview of the variation in methodologies between companies.

**Table 1.**
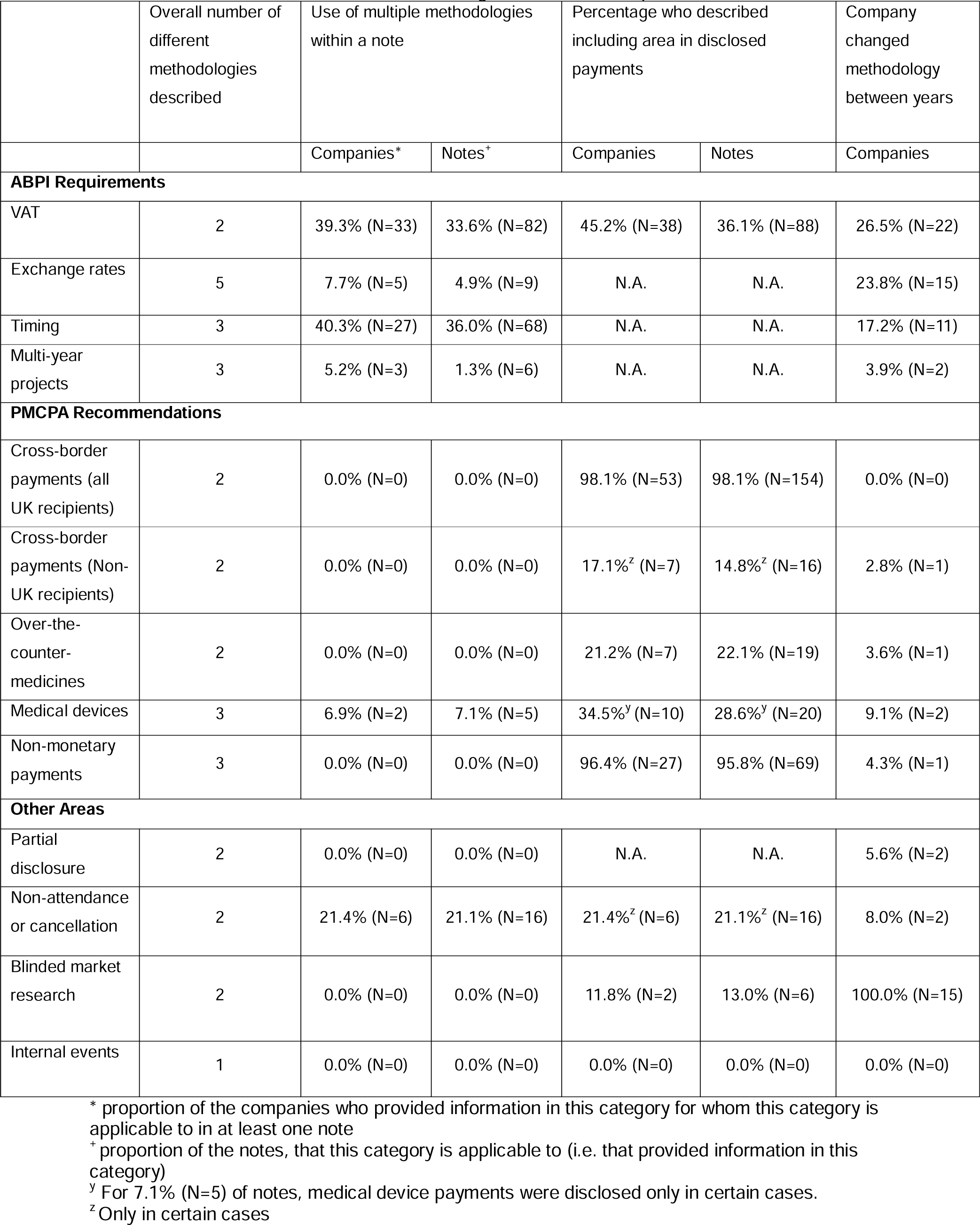
Overview of variation in methodologies for various aspects of disclosure.

Of the 244 notes with clear information on **VAT management**, 36.1% (N=88) included VAT, 30.3% (N=74) excluded VAT, and 33.6% (N=82) had multiple rules for VAT management. (Details of methodologies, with percentages and frequencies for companies, instead of notes, can be found in Appendix, eBox 2.)

Of the 182 notes with clear information on **exchange rate** methodology, 95.1% (N=173) reported one of five methodologies: 1) the exchange rate on date of payment (49.5%, N=90), 2) the average rate for the respective year (11.5%, N=21), 3) the average rate for the respective month (17.6%, N=32), 4) the exchange rate at another time e.g. date of entry (4.9%, N=9), or 5) the exact exchange rate was given (11.5%, N=21). For 4.9% (N=9) of notes they provided different rules depending on the circumstances.

Of the 189 notes with a clear methodology on how the **timing** of payments was handled, there were two methodologies: 1) 53.4% (N=101) used payment date for disclosure, and 2) 10.6% (N=20) used event date. For 36.0% (N=68), multiple rules were used i.e. in certain cases it was the payment date and in other cases it was the event date.

Of the 150 notes with a clear methodology on management of **multi-year projects**, 72.7% (N=109) assigned each payment in the multi-year contract to that payment’s respective year (this also included notes that assigned each payment in the multi-year contract to the relevant invoice’s respective year), 18.7% (N=28) assigned the payment to the year the ‘transfer of value’ occurred (e.g. the year the respective event took place), 4.7% (N=7) assigned the total value to the year in which the last payment was made, and 1.3% (N=6) used multiple rules.

### 3.3 Methodologies for PMCPA Recommendations

For PMCPA recommendations, there was a mix of methodologies. Of the 157 notes that provided clear information on their methodology for **cross-border payments to UK registered recipients,** for 98.1% (N=154), payments to UK registered recipients by a company’s affiliate outside the UK, and/or for activities outside the UK, were disclosed on Disclosure UK. For notes from Baxter International, payments to UK based HCPs/HCOs from the company’s US affiliate were not disclosed on Disclosure UK.

Of the 108 notes that provided clear information on their methodology for **cross-border payments to recipients registered outside the UK,** for 14.8% (N=16), payments to HCPs/HCOs based outside the UK were included in the UK disclosure system. In four notes this only applied to countries where the company did not have an affiliate. For 85.2% (N=92) of notes, it was explicitly stated that HCPs/HCOs based outside the UK were not included in UK disclosures.

Of the 86 notes that provided clear information on payments related to **over-the-counter-medicines**, 22.1% (N=19) stated they included them in their disclosure, whereas 47.7% (N=41) explicitly excluded them. For 30.2% (N=26) of notes, we inferred from their definition of includable payments as related to ‘prescription only medicines’ that payments related to over-the-counter-medicines were excluded.

Of the 70 notes that provided clear information on payments related to **medical devices**, 28.6% (N=20) included such payments in their disclosure, whereas 30.0% (N=21) explicitly excluded them. For 34.3% (N=24) of notes, we inferred from their definition of includable payments as related to ‘prescription only medicines’ that payments related to medical devices were excluded. For 7.1% (N=5) of notes, medical device payments were disclosed in certain cases, for example, for devices with active pharmacological ingredients.

Of the 72 notes that provided clear information on quantification of **non-monetary payments**, 50.0% (N=36) described quantifying non-monetary payments but did not detail a calculation method, whereas 45.8% (N=33) detailed a calculation method. In the case of Leo Pharma, their three notes described areas of ‘non-financial support […] that cannot be assigned a monetary value.’

### 3.4 Changes in Methodologies

Some companies changed methodologies between years. The ABPI requirements with the highest percentage of companies changing methodology was VAT; 26.5% (N=22) of companies who provided clear information in two or more of the included years changed methodology at least once. The PMCPA recommendation with the highest percentage of companies changing methodology was cross-border payments; 10.8% (N=4) of companies who provided clear information in two or more of the included years, changed methodology at least once. An overview of percentages of companies who changed methodology between years is in Table 1.

### 3.5 Methodologies for Other Areas

Of the 109 notes that provided clear information on **partial disclosure**, the practice of allowing individual HCPs to anonymise some payments they have received while consenting to being identified for other payments, 10.1% (N=11) permitted this practice, whereas 89.9% (N=98) did not.

Of the 76 notes that provided clear information on their methodology for **non-attendance or cancellation** (when a HCP cancelled or did not attend an event), 78.9% (N=60) did not disclose payments for these events, whereas 21.1% (N=16) of notes outlined that these payments were disclosed. However, this was only in certain cases, for example, if travel costs were paid by the HCP and reimbursed by the company, but the HCP did not take the journey.

Of the 46 notes that provided clear information on payments related to **blinded market research**, 87.0% (N=40) excluded these payments and 13.0% (N=6) included the payments but reported them in aggregate to ensure recipients remained anonymous.

Of the 36 notes that provided clear information on management of HCP attendance at **internal events,** for 100.0% (N=36) of notes, no proportion of the internal costs associated with these events were disclosed as payments to HCPs.

There were several different cases of payment areas **excluded** from disclosure, without any permission in ABPI guidance, details of which are below. Alimera Sciences outlined in one note that the provision of free stock to a HCP/HCO was not disclosed. Allergan, in all notes, outlined that ‘confidentiality clauses within contracts with HCOs may prohibit Allergan from disclosing the transfer of value.’ Finally, AstraZeneca and Bayer in all their notes, did not disclose administration fees associated with payments to HCPs/HCOs.

Some companies **included** other payment areas in disclosures that were not required to be included by the ABPI. For example, 1.9% (N=5) of notes from 3.3% (N=3) of companies mentioned including subsistence, at least partially. Also, 5.6% (N=15) of notes from 6.7% (N=6) of companies stated that they disclosed payments to retired HCPs.

Some companies reported payment areas in **aggregate** which were not explicitly permitted to be reported in aggregate by the ABPI. In relation to HCOs, 4.8% (N=13) of notes mentioned instances when a payment to a HCO would be reported in aggregate, for example if the HCO revokes consent. In 4.1% (N=11) of notes, payments were aggregated temporarily if the stated recipient disputed the payment amount or disputed whether the payment was given to them at all. For three notes, from two companies (Chugai and Novartis), payments to recipients who had since died were aggregated. For all notes from Swedish Orphan Biovitrum, payments ‘related to commercially sensitive information’ were aggregated. For two notes from Bristol Myers Squibb, payments were aggregated when ‘the name of a HCO contains the name of a HCP who is also the sole director of the HCO’. For all notes from Thea Pharmaceuticals no consent was obtained from HCPs and all HCP payments were aggregated.

## 4. Discussion

Drawing on methodological notes from three years, this study provides granular insight into shortcomings in how drug companies in the UK apply self-regulation in practice. The study shows that many companies did not adhere to ABPI requirements or PMCPA recommendations in relation to provision of information in their methodological notes on how key areas such as VAT are handled. For example, only three of the 90 companies analysed provided clear information for all PMCPA recommendations and ABPI regulations in all their notes. The persistent non-adherence to ABPI requirements over time indicates a lack of effective enforcement. The study also revealed differences in disclosure methodologies between companies, as well as individual companies changing methodologies between years. Overall, the findings demonstrate the limits of pharmaceutical industry self-regulation in relation to transparency, despite ABPI rhetoric about the importance of transparency.^42^

One key finding is that data published on Disclosure UK likely differs significantly from the picture developed when considering the methodological notes. For example, 36% of notes described including VAT when reporting payments, 30% excluded it, and 34% had different rules depending on the circumstances. A similar mix of VAT methodologies was found in Irish methodological notes.^18^ This undermines journalistic investigations examining COIs using Disclosure UK,^25,43^ as the methodological notes are unlikely to have been considered. Also, in important areas for interpreting payment disclosures, many methodological notes did not provide information. For example, for multi-year contracts, an area that the ABPI require notes to cover, 33% of notes did not provide clear information. These issues highlight major barriers to accurately mapping the financial links between pharmaceutical companies and healthcare. We also found potential breaches of the ABPI code; 5% of notes mentioned instances when a payment to a HCO might be reported in aggregate. The same figure was found in the Irish study of methodological notes.^18^ Also, in one methodological note, Thea pharmaceuticals described not obtaining consent from HCPs and aggregating all HCP payments. Given that the UK disclosure system is considered the most accessible self-regulated European system,^22,27^ similar issues likely exist with methodological notes in other EFPIA pharmaceutical markets (e.g. Spain).

This study adds to the literature showing major shortcomings of self-regulated payment disclosure systems from both the pharmaceutical^1,18,22,44,45^ and medical device^46^ industries. This study’s findings of opacity in several areas corresponds with issues outlined in other studies of disclosure systems, such as non-disclosure of certain payment areas,^27^ and unnecessary aggregation of recipients.^18,46^ Overall, it reinforces the conclusions of the 2021 analysis of European disclosure systems, that “self-regulation cannot address ‘the issues of perceived conflict of interest’, as promised by EFPIA.”^22^ The shortcomings of self-regulation found here reflect issues with self-regulation generally, which can be seen in other areas of the pharmaceutical industry^47,48^ and other industries including alcohol^49^ and nutrition.^50^ For example, a 2016 systematic review found that violations of self-regulation guidelines for alcohol marketing were ‘highly prevalent.’^49^

### 4.1 Recommendations and Future Research

The findings from this study and others,^22,27^ raise questions about the effectiveness of self-regulation as a governance tool.^51^ A mandatory system of pharmaceutical industry payment disclosure, similar to the US or French systems^15–17^ and developed independently of industry, should be considered in the UK. Depending on details of design and implementation,^22,45^ this would likely result in more reliable and valid disclosures. However, there are shortcomings in some mandatory systems. For example, the US Open Payments system does not appear to require methodological note publication.^52^ The UK government recently conducted a consultation in this area, with a mandatory system and an expanded self-regulatory system both under consideration.^26^ However, some have argued that the proposals may reduce transparency compared to the status quo.^24^

If states choose not to mandate transparency, then improvements to the self-regulatory system should be considered. Firstly, EFPIA and by extension, the ABPI, should standardise disclosure rules. For example, VAT should be included in all disclosures, where applicable, as is required by the European medical device industry.^46^ For other areas, where different methodologies are permissible due to different business practices (e.g. exchange rates) adherence to the EFPIA template methodological note (Appendix, eBox 3) would improve interpretability. However, a more user-friendly approach might be to integrate methodology information with the database itself. Another recommendation for the ABPI is to publish a single file in an analysable format detailing companies’ methodologies. Though this study is primarily focussed on transparency, we caution against a disproportionate focus on transparency. Transparency itself has limited effects on addressing issues associated with COIs.^21,53^ Attention should be directed towards managing, reducing and, in cases, eliminating COIs.

In relation to future research, a study combining information from the methodological notes with payment disclosure data would provide information on how much the value of payments would change if payment rules were applied consistently. Other research could involve a comparison of disclosure methodologies in other countries and in the medical device industry.

### 4.2 Strengths and Limitations

This study’s primary strength is that, to our knowledge, this is the most extensive analysis of industry methodological notes to date. Secondly, by sharing the coded data for methodological notes, this could be an important resource for anyone conducting future analysis using Disclosure UK data to adjust for methodological differences. There were also limitations. Data was not analysed for the years 2020-2022, which were available at time of submission, due to the large workload involved in extracting and analysing data.

Furthermore, the exclusion of companies who only reported payments in two of the three included years was a limitation. However, companies reporting payments in all three years represent 95% of Disclosure UK payments in terms of value.

### 4.3 Conclusion

This study shows major deficiencies in the self-regulated pharmaceutical industry payment disclosure system operating in the UK. Specifically, we identified that many companies, in their methodological notes, did not provide information required by the ABPI, and amongst those that did provide information there was large variation in methodologies. These findings point to limitations of pharmaceutical industry self-regulation. To effectively regulate this area, a state mandated system of pharmaceutical industry payment disclosure could be introduced, addressing the issues identified in this study and others.

## Data Availability

Data are available online at: Larkin, J. (2024). Disclosure UK Methodological Notes Data 2015, 2017, 2019 [Data set]. Zenodo.

https://doi.org/10.5281/zenodo.10953205

