## Appendix A for "Drug company methodologies used for reporting in the UK pharmaceutical industry payment transparency database between 2015-2019: A content analysis"

eTable 1 Examples of the categorisation

|  | Clear information provided | Information provided but unclear |
| --- | --- | --- |
| VAT | ‘All values reported are exclusive of VAT’ | ‘VAT is only paid where the recipient is VAT registered and has provided a bone fide VAT invoice.’ |
| Exchange rates | ‘Where a transfer of value is made in a currency other than GBP Alexion will convert at HMRC 2017 average exchange rates.’ | ‘The Disclosure Report will show transfers of value made in GDP. Conversion from local currency to GDP will be carried out at the time of the generation of the report.’ |
| Timing | ‘This disclosure submission includes those payments and ToVs that are subject to Britannia disclosure  obligations covering the period 1 January 2019 to 31 December 2019.’ | ‘The information provided has been derived from invoices coded into the financial systems for the relevant period.’ |
| Multi-year projects | ‘For multi year contracts, payments are disclosed in the year corresponding to the date of each payment’ | ‘For multiyear contracts, disclosure only includes ToVs applicable during the reporting period (1 January 2015 and 31 December 2015).’ |
| Cross-border payments | ‘All payments worldwide are declared as Diurnal is headquartered in the UK’ | ‘Cross-border payments: All payments made to UK HCPs or HCOs from outside the UK are tracked and managed centrally by our European team. All information is contained within our financial system outlining full payments made including travel and associated expenses. Contracts are also shared with the UK office containing full payment details for each engagement. The PTC Finance Team are able to track all external payments made to HCPs and HCOs within the UK.’ |
| Over-the-counter medicines | ‘In accordance with the EFPIA Disclosure Code and the ABPI Code of Practice, Novo Nordisk does not disclose the following items: i) over-the-counter medicines’ | No entries were categorised as *Information provided but unclear* for over-the-counter medicines |
| Medical Devices | ‘As long as ToVs are not exclusively connected to over-the-counter medicines or medical devices – which are not in scope of the EFPIA Disclosure Code - Bayer will disclose such ToVs in full.’ | No entries were categorised as *Information provided but unclear* for medical devices |
| Non-Monetary Payments | ‘If a ‘donation in kind’ is provided to a HCO e.g. Amgen staff time, a monetary value will be attributed to the ‘in kind’ donation for the purpose of disclosure.’ | ‘How should Member Companies handle the more intangible Transfers of Value relating to medical publications support which could be “in kind” transfers of value or indirect support? Answer: This answer need to be review by the drafting group’ |
| Partial disclosure | ‘Partial Consent: Almirall Ltd. collects consent on a per-activity basis. Where recipients of transfers of value have decided to disclose as an individual in some activities and aggregate in others, the amount attributable to all transfers is disclosed on an aggregate basis’ | ‘If a HCP revokes the consent to disclose under the Disclosure Code, Baxter will aggregate the data for the future but not retrospectively. Any consequences of withdrawal of consents shall be defined in the relevant Agreement.’ |
| Non-attendance or cancellation | ‘In case of cancellation the ToV is not included in the disclosure. In case of partial attendance the ToV is included in the disclosure’ | No entries were categorised as *Information provided but unclear* for medical devices |
| Blinded market research | ‘ToV paid as part of a market research study where Eisai does not know the identity of the participating HCPs or HCOs will be disclosed on a aggregated basis. if available.’ | No entries were categorised as *Information provided but unclear* for medical devices |
| Internal events | ‘Costs for internal Events such as rent for space, technical expenses or equipment hire will not be disclosed within the Disclosure Report.’ | No entries were categorised as *Information provided but unclear* for medical devices |

eTable 2. Whether information was provided for key methodological areas and whether policies changed between years

|  | Clear information provided | | Information provided but unclear | | Information not provided | | Not applicable | | Company changed methodology between years^x^ |
| --- | --- | --- | --- | --- | --- | --- | --- | --- | --- |
|  | Com* | Note^+^ | Com | Note | Com | Note | Com | Note |  |
| **ABPI Requirements** | | | | | | | | | |
| VAT | 93.3%  (N=84) | 90.7%  (N=244) | 3.3% (N=3) | 2.6% (N=7) | 10.0% (N=9) | 6.7% (N=18) | 0.0% (N=0) | 0.0% (N=0) | 26.5%  (N=22) |
| Exchange rates | 71.1%  (N=64) | 67.7%  (N=182) | 24.4% (N=22) | 20.1% (N=54) | 8.9% (N=8) | 7.1% (N=19) | 6.7% (N=6) | 5.2% (N=14) | 23.8%  (N=15) |
| Timing | 74.4%  (N=67) | 70.6%  (N=190) | 10.0% (N=9) | 8.2% (N=22) | 24.4% (N=22) | 21.2% (N=57) | 0.0% (N=0) | 0.0% (N=0) | 17.2%  (N=11) |
| Multi-year projects | 64.4%  (N=58) | 55.8%  (N=150) | 1.1% (N=1) | 0.4% (N=1) | 41.1% (N=37) | 32.7% (N=88) | 14.4% (N=13) | 11.2% (N=30) | 9.8%  (N=5) |
| **PMCPA Recommendations** | | | | | | | | | |
| Cross-border payments | 66.7%  (N=60) | 63.6%  (N=171) | 6.7% (N=6) | 5.6% (N=15) | 31.1% (N=28) | 26.4% (N=71) | 5.6% (N=5) | 4.5% (N=12) | 10.8%  (N=4) |
| Over-the-counter medicines | 36.7% (N=33) | 32.0%  (N=86) | 0.0% (N=0) | 0.0% (N=0) | 58.9% (N=53) | 51.3% (N=138) | 16.7% (N=15) | 16.7% (N=45) | 3.6%  (N=1) |
| Medical devices | 32.2%  (N=29) | 26.0% (N=70) | 0.0% (N=0) | 0.0% (N=0) | 70.0% (N=63) | 61.7% (N=166) | 15.6% (N=14) | 12.3% (N=33) | 9.1%  (N=2) |
| Non-monetary payments | 31.1%  (N=28) | 26.8% (N=72) | 1.1% (N=1) | 0.4% (N=1) | 66.7% (N=60) | 61.7% (N=166) | 15.6% (N=14) | 11.2% (N=30) | 4.3%  (N=1) |
| **Other Areas of Interest** | | | | | | | | | |
| Partial disclosure | 44.4%  (N=40) | 40.5%  (N=109) | 1.1% (N=1) | 1.1% (N=3) | 62.2% (N=56) | 58.4% (N=157) | 0.0% (N=0) | 0.0% (N=0) | 5.6%  (N=2) |
| Non-attendance or cancellation | 31.1%  (N=28) | 28.3%  (N=76) | 0.0% (N=0) | 0.0% (N=0) | 74.4% (N=67) | 71.7% (N=193) | 0.0% (N=0) | 0.0% (N=0) | 8.0%  (N=2) |
| Blinded market research | 18.9%  (N=17) | 17.1%  (N=46) | 0.0% (N=0) | 0.0% (N=0) | 84.4% (N=76) | 82.9% (N=223) | 0.0% (N=0) | 0.0% (N=0) | 100.0%  (N=15) |
| Internal events | 17.8%  (N=16) | 13.4%  (N=36) | 0.0% (N=0) | 0.0% (N=0) | 90% (N=81) | 86.6% (N=233) | 0.0% (N=0) | 0.0% (N=0) | 0.0%  (N=0) |

* proportion of the 90 companies for whom this category is applicable to in at least one note

^+^ proportion of the 269 notes this category is applicable to

^x^ Proportion of companies who provided information in at least two years who had different methodologies in at least two of the three analysed years for the given area

eBox 1. Definition of HCPs, HCOs, ORDMs, and payment areas according to the ABPI disclosure code^1^

‘Other relevant decision maker ‘is someone with an NHS role who could influence in any way the administration, consumption, prescription, purchase, recommendation, sale, supply or use of any medicine but who is not a health professional

'Health professional' includes any member of the medical, dental, pharmacy or nursing profession and any other person who in the course of their professional activities may administer, prescribe, purchase, recommend or supply a medicine. In relation to the annual disclosure of transfers of value (Clause 28), the term also includes any employee of a pharmaceutical company whose primary occupation is that of a practising health professional

'Healthcare organisation' means either a healthcare, medical or scientific association or organisation such as a hospital, clinic, foundation, university or other teaching institution or learned society whose business address, place of incorporation or primary place of operation is in Europe or an organisation through which one or more health professionals or other relevant decision makers provide services.

‘Contracted services’: Health professionals, other relevant decision makers or their employers on their behalf, healthcare organisations, patient organisations, individuals representing patient

organisations, and members of the public, including patients and journalists, may be used as consultants and advisors, whether in groups or individually, for services such as speaking at and chairing meetings, involvement in medical/ scientific studies, clinical trials or training services, writing articles and/or publications, participation at advisory board meetings, and participation in market research where such participation may involve remuneration and/or hospitality.

‘Contributions to cost related to events’ means providing or covering the costs of travel, accommodation and/or registration fees to support the attendance of an individual to an event organised or created by a company and/or independent organisation. When providing sponsorship of events/meetings to organisations, associations etc such contributions may include costs for subsistence (food and drink).

‘Donations and grants' collectively mean providing funds, benefits-in-kind or services freely given for the purpose of supporting healthcare, scientific research or education, with no consequent obligation on the recipient organisation, institution and the like to provide goods or services to the benefit of the pharmaceutical company in return. Donations and grants to individuals are prohibited. In general, donations are physical items, services or benefitsin-kind which may be offered or requested. Grants are the provision of funds.

‘Joint working’ is defined as situations where, for the benefit of patients, one or more pharmaceutical companies and the NHS pool skills, experience and/or resources for the joint development and implementation of patient centred projects and share a commitment for successful delivery. Each party must make a significant contribution and the outcomes must be measured.

‘Research and development’ means, for the purposes of disclosure, transfers of value to health professionals or healthcare organisations related to the planning or conduct of: i. non-clinical studies (as defined in the OECD Principles on Good Laboratory Practice) ii. clinical trials (as defined in Regulation 536/2014) iii. non-interventional studies that are prospective in nature and that involve the collection of patient data from or on behalf of individual or groups of health professionals specifically for the study

eBox 2. Details of methodologies broken down by company

### Methodologies for ABPI Requirements

Of the 84 companies that provided clear information on **VAT management** in at least one year, 45.2% (N=38) included VAT in at least one year, 41.7% (N=35) excluded VAT in at least one year, and 39.3% (N=33) had multiple rules for VAT in at least one year.

Of the 65 companies which provided details on an **exchange rate** methodology in at least one year, companies reported one of five methodologies in at least one year: 1) the exchange rate on date of payment (60.0%, N=39), 2) the average rate for the respective year (13.8%, N=9), 3) the average rate for the respective month (20.0%, N=13), 4) the rate at another time such as the date of entry (7.7%, N=5), or 5) the exact exchange rate was given (13.8%, N=9). For 7.7% (N=5) of companies provided different rules depending on the circumstances.

Of the 67 companies which provided details on a **timing** methodology in at least one year, companies reported one of two methodologies in at least one year: 1) 64.2% (N=43) of companies reported using the payment date, and 2) 11.9% (N=8) reported using event date. For 40.3% (N=27) of companies, in at least one of their notes multiple rules were used i.e. in certain cases it is the date of payment and in other cases it is the event date.

Of the 58 companies that provided a clear methodology on how **multi-year projects** were managed in at least one year, 72.4% (N=42) of companies described assigning each payment in the multi-year contract to that payment’s respective year (this also included notes that described assigning each payment in the multi-year contract to that invoice’s respective year), 5.2% (N=3) assigned the total value to the year in which the last payment was made, 20.7% (N=12) of notes assigned the payment to the year the transfer of value occurred (this could occur in a single year or there could be several transfers of value in several different years), and 5.2% (N=3) used multiple rules.

### Methodologies for PMCPA Recommendations

Of the 54 companies that provided clear information on management of **cross-border payments to UK registered recipients** in at least one note, for 98.1% (N=53) of companies, payments to UK registered recipients by a company’s affiliate outside the UK, and/or for activities outside the UK, were disclosed on disclosure UK. For all three notes from Baxter international, payments to UK based HCPs/HCOs from the company’s US affiliate were not included.

Of the 41 companies that provided clear information on management of **cross-border payments to recipients** **registered outside the UK** in at least one note, for 17.1% (N=7) of companies, payments by the respective company, to HCPs/HCOs based outside the UK were included in the UK disclosure system. Though it should be noted that in some cases this only applied to countries where the company did not have an affiliate. For 85.4% (N=35) of companies, it was explicitly stated that HCPs/HCOs based outside the UK were not included in the UK disclosure system.

Of the 33 companies that provided details in at least one note of a methodology for payments related to **over-the-counter medicines**, 21.2% (N=7) stated they included them in their disclosure, whereas 54.5% (N=18) explicitly excluded them. For 33.3% (N=11) of notes, it can be inferred from their definition of includable payments as related to ‘prescription only medicines’ that payments related to over-the-counter medicines were excluded.

Of the 29 companies that provided details in at least one note of a methodology for payments related to **medical devices**, 27.6% (N=8) included such payments in their disclosure, 34.5% (N=10) explicitly excluded them. For 37.9% (N=11) of companies, in at least one of their notes it can be inferred from their definition of includable payments as related to ‘prescription only medicines’ that payments related to medical devices were excluded. For 6.9% (N=2) of companies, in at least one of their notes medical device payments were partially disclosed, for example, for devices with active pharmacological ingredients.

Of the 28 companies that provided clear details in at least one note of a methodology on how **non-monetary payments** were quantified, 53.6% (N=15) described quantifying non-monetary payments but did not provide details of a calculation method, 46.4% (N=13) did provide details of a calculation method. In the case of Leo Pharma, all three of their notes describe several areas of ‘non-financial support […] that cannot be assigned a monetary value.’

### Methodologies for Other Areas

Of the 40 companies that provided, in at least one note, a clear methodology on **partial disclosure**, the practice of allowing individual HCPs to anonymise some of the payments they have received while consenting to being identified for other payments, this was permitted in Y% (N=Y) of companies. For Y% (N=Y) of companies did not permit partial disclosure.

Of the 28 companies that provided, in at least one note, a clear methodology on **non-attendance or cancellation** (when a HCP cancelled or did not show up to an event): 85.7% (N=24) of companies did not include these in their disclosure. Whereas 21.4% (N=6) did include such payments in their disclosure but only in certain cases, for example, if travel costs were paid by the HCP and reimbursed by the company, but the HCP did not take the journey.

Of the 17 companies that provided, in at least one note, information on how they managed **blinded market research**, 88.2% (N=15) excluded these payments and 11.8% (N=2) included the payments but reported them in aggregate to ensure recipients remained anonymous.

Of the 16 companies that provided information on how they managed HCPs attendance at **internal events,** for 100.0% (N=16) of companies, no proportion of the internal costs associated with these events were disclosed as indirect payments to HCPs.

eBox 3. EFPIA methodological note template^1^

**HCP/HCO Disclosure**

**Methodological note structure’s template**

**Introduction**

In 2014, by introducing the disclosure requirements, EFPIA and its members have demonstrated their commitments to self-regulation but also the legitimacy of the interactions with HCPs, and HCOs.

Member Companies and companies that are members of Member Associations are required to disclose transfers of value^[1]^ made to HCPs and HCOs. This disclosure includes, by HCP or HCO, the total amounts of value transferred, by type of activity.

The disclosure must be aligned with local laws and regulations. The methodological note is required for disclosure based on the EFPIA Code requirements as transposed in national codes provisions.

The methodological note is usually made for a specific country; if this is the case, please specify the country concerned.

In addition to the methodological note in the local language, it is recommended to have an English version.

**Definitions**

- Recipients (Type of recipients included – If needed, include the definition of Healthcare Professional at national level – Treatment of retired and deceased HCPs)
- Kind of ToVs (donations and grants – contribution to costs of events – fees for services and consultancy – R&D^[2]^ – others)

**Disclosure’s scope**

- Products concerned (Products included in the disclosure report: POM, others)
- Company concerned (affiliate – merger – company rebrand)
- Excluded ToVs
- ToVs date
- Direct ToVs
- Indirect ToVs
- Non-monetary ToVs
- ToVs in case of partial attendances or cancellation and refund
- Cross-border activities
- R&D
- Voluntary disclosure (anything discloses beyond the national Code)

**Specific considerations**

- Country unique identifier (if needed, specify which identifier is used and for which purpose)
- Self-incorporated HCP (depending on the local legislation, qualified as individual or company)
- Multi-year agreements
- Country specificities
- Quality Checks (optional for pre-disclosure)

**Data protection legal basis**

- Consent collection (inc. consent withdrawal)
  - Partial consent
- Legitimate interest (inc. balancing test, right to object)

**Form of Disclosure**

- Date of publication
- Disclosure platform
- Disclosure language

**Disclosure financial data**

- Currency (local or if not, specify the exchange rate)
- VAT included or excluded
- Calculation rules (e.g., in-kind ToVs)

**Additional information**

1. Transfers of Value are direct and indirect payment, whether in cash, in kind or otherwise or reimbursement provided to HCPs and HCOs.
2. Cf. EFPIA’s R&D best practice guidance: <https://efpia.box.com/s/nms7cod07bbkwwjenr3kcyerdk17k4xw>
